# Death, Inequality, and the Pandemic in the Nation’s Capital

**DOI:** 10.1101/2022.12.02.22283039

**Authors:** Maria L. Alva, Srujana S. Illa, Jaren Haber

**Affiliations:** Massive Data Institute, McCourt School of Public Policy, Georgetown University, Washington DC, USA; Quantitative Social Science, Dartmouth College

## Abstract

Abrupt changes in mortality rates and life expectancy allow us to understand how shocks like COVID-19 can exacerbate health inequalities across groups. We look at Washington, D.C., a major city with a diverse population and long-standing socio-economic divisions, to describe the all-cause mortality trends from 2015 to 2021 by age, sex, race, and ward of residence. We report differences in cause-specific mortality pre- and post-COVID-19 outbreak and estimate the Years of Life Lost (YLL) attributable to COVID-19. We compute death rates using information from death certificates and the Census, and we calculate YLL using the life table approach, comparing the life expectancy of people with and without COVID-19. We find that in 2020 and 2021, there were respectively 1,128 and 629 excess deaths (158 per 100K and 94 per 100K) compared to the annual average over the previous five years, and 689 and 363 deaths in 2020 and 2021, respectively (97 per 100K and 54 per 100K) listing COVID-19 as a cause of death. Death rates in 2020 and 2021, compared to the five previous years, were higher for men than women by about 12pp and 5pp and occurred almost entirely among residents 45 and older. Excess deaths between 2020 and 2021 were higher for Black and Hispanic residents by about 286 and 97 per 100K, respectively—with the highest proportional increase (almost twofold) for Hispanics in 2020. YLL was highest for Hispanic males and lowest for White females.

**Significance Statement:** The leading causes of death in DC were historically heart disease and cancer. In 2020, the leading cause of death was COVID. Accidental and violent deaths increased dramatically and disproportionately by race. Racial disparities in COVID and non-COVID deaths indicate that these correlate with socioeconomic conditions.

Life expectancy in the United States decreased for the first time in 2020 due to COVID-19. In the nation’s capital, the decline in life expectancy was more significant for Hispanic and non-Hispanic Black than White people, widening the already large difference in life expectancy among these groups.

## Introduction

As of December 2022, the CDC has reported more than 1.1M deaths from COVID-19 in the US (1). Herein, we quantify how the pandemic has impacted health outcomes in Washington, DC, a city of approximately 712,816 people with over a thousand documented COVID-19 deaths across the first two years of the pandemic (1, 2).

Because DC is a socioeconomically and racially diverse city, the data describing all-cause mortality, cause-specific mortality, and life expectancy provides an informative case study to understand the extent to which the pandemic has widened pre-existing gaps across sociodemographic groups. Washington, DC, has been consistently listed in the top 10 cities for inequality in the past five decades, together with New York, Chicago, San Francisco, and Los Angeles (3). This paper quantifies the mortality burden during COVID-19 for a large, economically unequal city like the nation’s capital.

We primarily track all-cause mortality because monitoring and reporting COVID-19-deaths alone has been shown to be limiting. For example, COVID-19 might not have been listed as a cause of death in death certificates because people who died had not been diagnosed or died of other comorbidities and, therefore, under-reported. COVID-19 might have also impacted deaths by increasing mental health disorders, violence, substance abuse (4), and the risk of otherwise preventable deaths made more likely by reduced use of health care services (5). On the other hand, reductions in mobility and social distancing mandates might have prevented non-COVID deaths for some groups. For example, flu transmission decreased by 20 percent (6), potentially saving lives.

Given these complexities, tracking all-cause mortality allows us to account for changes possibly induced by policy and behavioral responses while avoiding potential measurement bias from focusing on COVID-19 mortality alone.

We also report changes in the documented causes of death pre-and post-pandemic to understand the indirect consequences of COVID-19 on other cause-specific deaths.

The contraction of healthcare consumption at the start of the pandemic (7) and the reductions in mobility under social distancing mandates might have impacted deaths in unforeseeable ways, leading to an increase in deaths that are typically preventable (4). In other words, we anticipate that COVID-19 might increase reported deaths from other causes, either because COVID-19 was under-reported in surveillance data, the pandemic derailed health services on potentially avoidable deaths, or violence and accidental deaths increased. We also anticipate that these excess deaths might not be equally borne across all groups; therefore, we report the demographic characteristics of those more likely to die from them. Historically, the leading causes of death in Washington, DC, have been heart disease, cancer, stroke, and diabetes (8), and the burden of these illnesses has been unequally distributed across DC’s wards (electoral districts), which are highly stratified along socio-economic and racial lines.

Finally, we compute Years of Life Lost (YLL). Mortality rates and YLL capture different facets of population health. Mortality rates capture the probability of dying in a given year. In contrast, YLL captures premature mortality in relation to how long a person is expected to live and therefore provides richer information than the mortality rate alone, because groups (e.g., sex and race strata) with vastly different life expectancies might have similar mortality rates. Decreases in life expectancy, for example, can have human capital and productivity consequences and intergenerational implications for social mobility, given the well-documented economic and emotional difficulties of single-parent families (9) and orphans (10).

## Data and Methods

As the primary source of information on mortality across socio-demographic groups, we use death certificates for 2015-2021 from the DC Department of Health’s Vital Records (11). Death certificates represent DC residents who passed away in DC (rather than elsewhere) and contain information on the deceased person’s name, date of birth, date of death, zip code of residence, race, age, sex, underlying causes, and manner of death.

For contextual demographic information on DC, we use the Census Bureau’s American Community Survey (ACS) annual estimates (12). To create ward-level demographic strata, we merge the residential zip codes in the DC death certificates with the wards’ longitude and latitude shape files (13) and ward-level demographic data (14).

### Mortality rates

We compute the all-cause mortality rate by demographic strata (i.e., age, race, sex, and ward) by combining death certificate counts with ACS data. As the ACS represents estimates based on annual data collected from a sample of housing units, these data present a margin of error (15). We report the ACS confidence intervals in Appendix A but do not account for the lower and upper bounds of the live population in any given year (denominator) in the mortality rate estimates, because deaths (numerator) represent population values rather than estimates as well as rare events (i.e., changes in the denominator estimates will not affect the overall rate). We also look at the changing demographics of all-cause mortality as well as the changing composition of causes of death between 2015-2019 (pre-pandemic) and 2020-2021.

### Years of Life Lost (YLL)

We estimate the impact of COVID-19 on YLL across demographic groups in DC by measuring the difference in life expectancy between having and not having COVID-19. To compute YLL we use 2020 (the most recently published) sex-and race-specific life tables for the US (16). We construct the COVID-specific life table for DC using all-cause mortality rates from CDC’s National Center for Health Statistics by single year of age and sex; DC-specific COVID prevalence by age, sex, and race (Appendix B); and DC-specific mortality risk by race (Appendix C) for those with and without COVID-19. We scale the all-cause mortality of those with COVID relative to those without. We compute a scale-up factor (Pharoah & Hollingworth, 1996), θu, ranging between *r* and 1, that incorporates the relative risk (*r*) and COVID prevalence (p) as shown here:

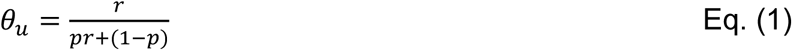

We compute the corresponding scale-down factor θd (Gray et al., 2010) for mortality for people without COVID as follows:

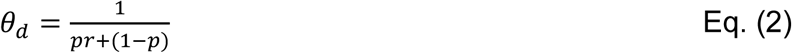

To compute *r*, we use the information on the cause of death (1 for COVID-19, 0 otherwise); to compute *p*, we use COVID-19 case counts between January 2020 and December 2021 for DC (17).

Existing CDC life tables estimate the probability of dying between a given age *x* and age *x*+1 (i.e., the number of individuals dying between age *x* and *x*+*t* divided by the number of individuals reaching exact age *x*). Using this information, we estimate the number of person-years lived, denoted as *L*_*x*_, between age *x* and *x*+*t*, as follows:

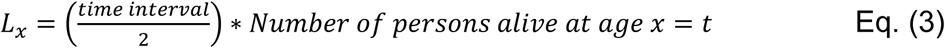

We calculate the total number of person-years that would be lived at a given age by cumulating the number of person-years lived from the oldest to the youngest age. We then calculate the average remaining lifetime (in years) for a person who survives a given age interval by dividing the total number of person-years lived from age *x* (*T*_*x*_) by the number of persons alive at age *x* (lx) (i.e., life expectancy = *T*_*x*_/*l*_*x*_). For deaths that occurred within the age interval *x* and *x*+*t*, the expected YLL equals the longest life expectancy for each cohort in the absence of COVID-19 minus the life expectancy with the condition.

## Results

### Mortality rates

When analyzing death counts by month, as in **Figure 1**, we observe that in 20 out of 24 months between 2020 and 2021, the death counts were significantly higher than the 2015-2019 five-year average. In particular, April, May, and December 2020, as well as January 2021, saw spikes of over 50% compared to the same months in previous years. DC recorded its first COVID case on March 7^th^ and its first COVID death on March 20^th^ (18, 19). The mayor of DC declared the state of emergency on March 11^th^ (20), the same day the World Health Organization announced COVID-19 a worldwide pandemic, and two days before COVID was declared a National Emergency (21).

**Figure 1.**
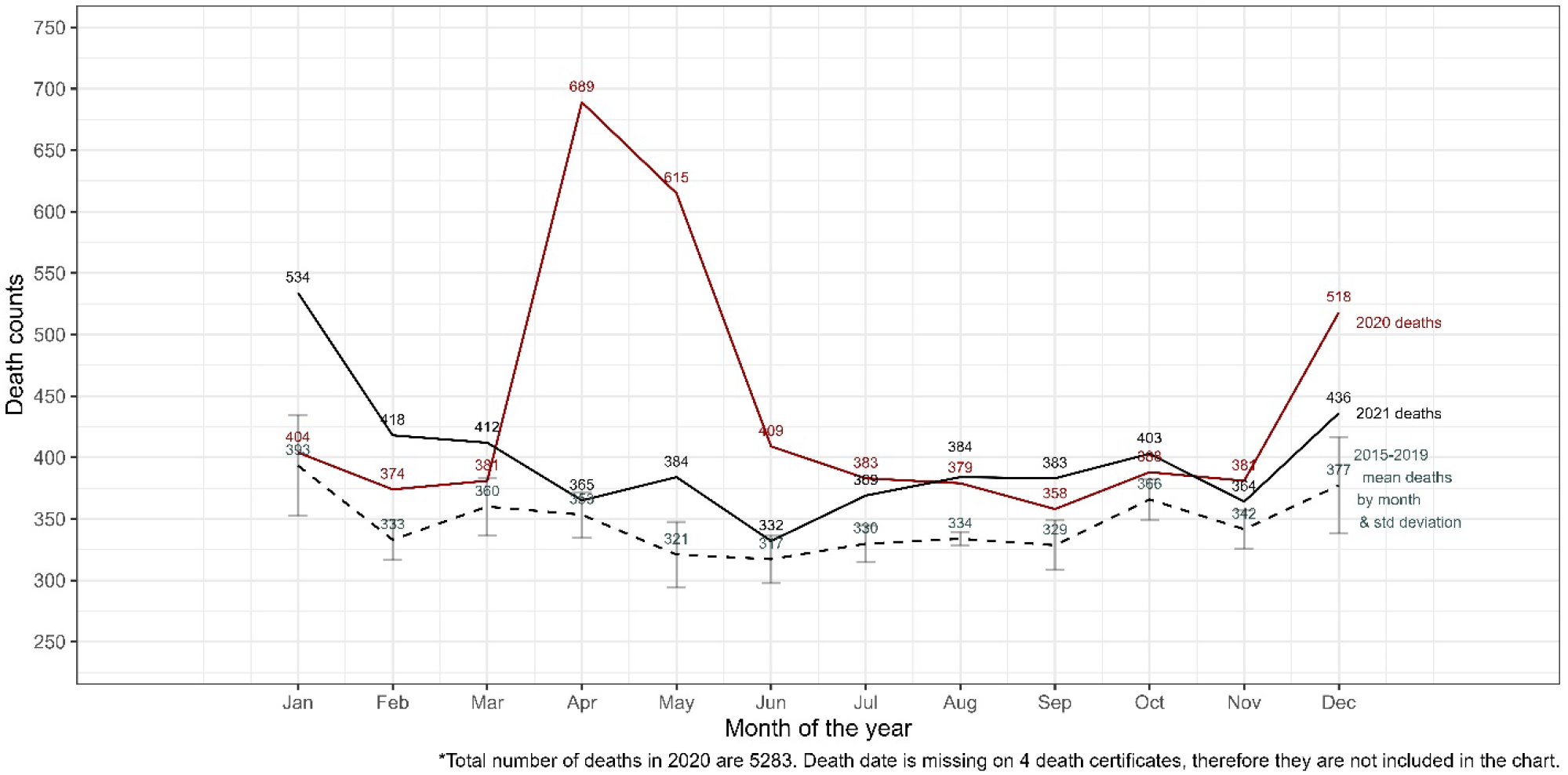
Monthly death counts in Washington, DC, far outpaced previous years in April, May, and December, 2020, and in January, 2021.

Schools, indoor venues, and non-essential businesses were closed for the next two months, and large gatherings were banned (22). Given what we have learned about transmissibility (23) and deaths, it is clear that the first cases happened at least one or two months earlier than officially recorded. Despite the stay-at-home orders, there were 689 deaths in April 2020 and 615 deaths in May 2020—a 95% and 92% increase compared to the 2015-2019 averages for those same months. The simultaneous spread of the Delta and Omicron variants led to the second peak in December 2020, following large-scale infection opportunities including the September 26th White House Rose Garden super-spreader event, Thanksgiving, and end-of-year religious gatherings (24).

Washington, DC, is a city of approximately 700,000 people (Appendix A) with a well-documented history of inequity (Appendix D). In 2020 and 2021 there were 5,283 and 4,784 deaths, respectively, which translates into 1,128 and 629 excess deaths compared to the previous five years’ average (2015-2019) after accounting for a population changes this translates to 158 and 94 excess deaths per 100K in 2020 and 2021, respectively. Almost all of these excess deaths were concentrated among residents aged 45 and over. Appendix E shows the age distribution of the mortality rate per 100,000 people in Washington, DC, from 2015-2021.

Figure 2 shows that the already large 2015-2019 mortality gap between the Non-Hispanic Black population and other racial groups increased in 2020 by a further 28% compared to 2019 and sustained at this level in 2021. Non-Hispanic Blacks represent the majority of deaths across all age groups, with 1,292 deaths per 100,000 in 2020 and 1,291 per 100,000 in 2021, six times the rate of the Non-Hispanic White population.

**Figure 2.**
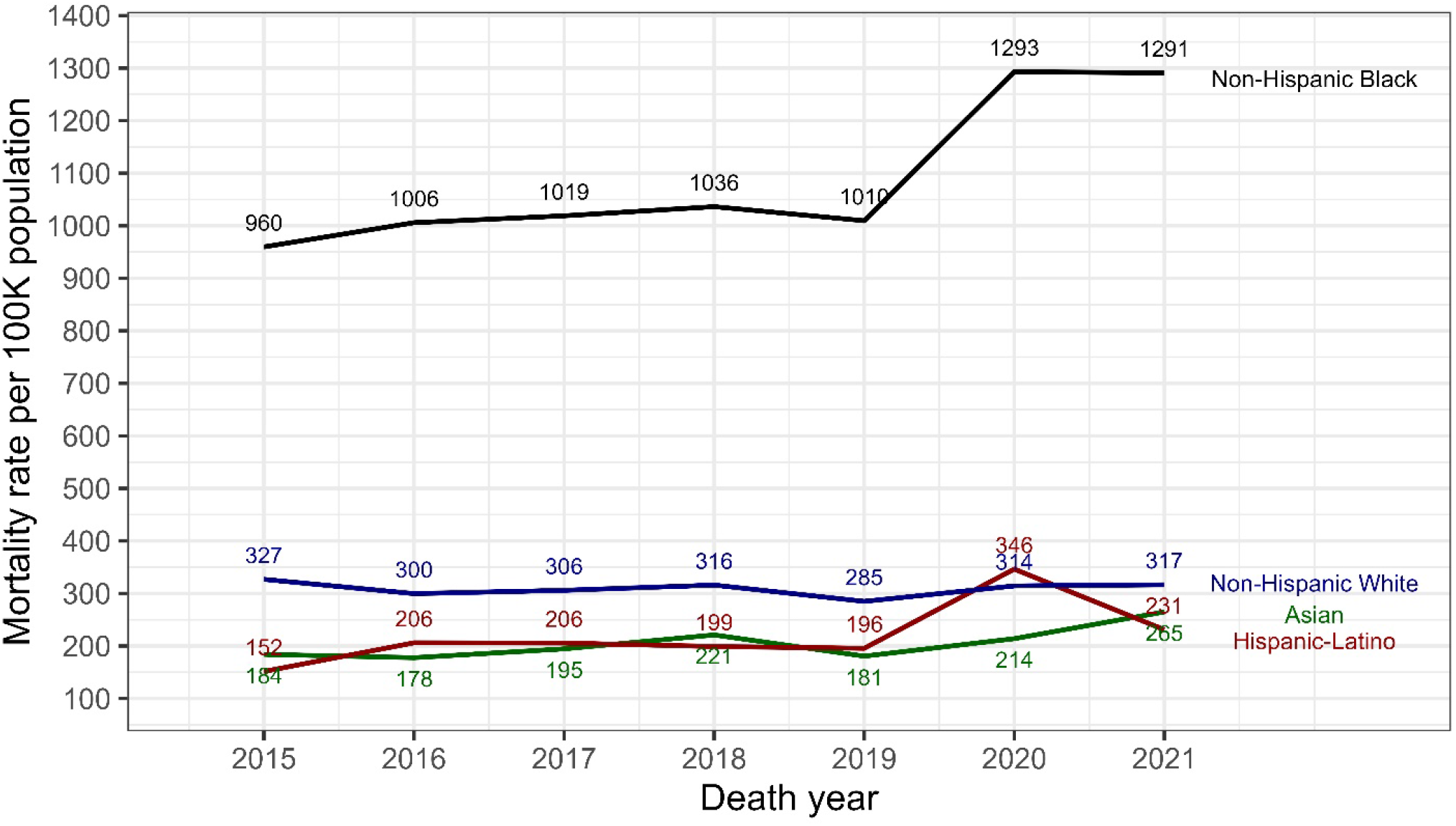
Existing racial disparities in DC mortality rates were further amplified in 2020 and 2021.

Hispanic/Latinos, on average a socioeconomically disadvantaged group with similar health risks as Blacks, have historically had a longer life expectancy than Black or Whites (a trend called the “Hispanic paradox” (25)). In 2020, however, Hispanics had the highest increase in mortality rates compared to all other groups, an almost twofold increase in mortality compared to the five previous years. In 2021, the Hispanic mortality rate decreased somewhat but did not drop to the pre-pandemic level. For context, DC’s population is 44% Non-Hispanic Black, 37% Non-Hispanic White, 11% Hispanic/Latino, and 4% Asian (see Appendix A for Census estimates). Over the past ten years, the Non-Hispanic Black population in DC has declined by 5.9 %, while the Non-Hispanic White and Hispanic/Latino population has increased by 2.5% and 2.2%, respectively (26).

Inequality in mortality rates has also widened along geographic and socio-economic dimensions, reflecting racial inequities and residential segregation. For planning and administrative purposes, Washington, DC, is divided into eight wards, each with approximately 88,000 residents. These wards are distinguished from one another by the racial/ethnic groups concentrated therein. The wards with the highest increases in mortality in 2020 relative to the 2015-2019 averages in decreasing order are wards 1 (45%), 7 (28%), 5 (25%), 4 (20%), 6 (18%), 8 (18%), 3 (<0.5%), 2 (<0.5%). Wards 2 and 3 are predominantly White, while wards 5, 6, 7, and 8 are mostly Black. Wards 1 and 4 have a relatively large (>20%) and growing Hispanic/Latino population and are also the most demographically diverse (27). Wards 5, 6, 7, and 8 are considered low-income communities. Wards 7 and 8, in particular, have unemployment rates over 20%. In contrast, wards 2 and 3 have unemployment rates markedly below the national average (28). In terms of health outcomes, wards 5, 6, 7, and 8 historically have also had the highest prevalence of chronic conditions in the district (8), and the mortality rate has been highest historically in Wards 5, 7, and 8. While mortality rates decreased in 2021 across all wards, only for wards 1, 2 and 3, rates reverted to pre-pandemic levels (**Appendix F**).

Figure 3 shows that the mortality gap by sex also increased in 2020 and 2021. The mortality rate of female residents was 652 deaths per 100,000 in 2020 and 644 per 100,000 in 2021, compared to the male mortality rate of 839 deaths per 100,000 and 790 per 100,000 in 2020 and 2021, respectively. This means a 17% and 16% increase in mortality for women and a 29% and 21% increase in mortality in mortality for men in 2020 and 2021, respectively, compared to the five-year, pre-pandemic average.

**Figure 3.**
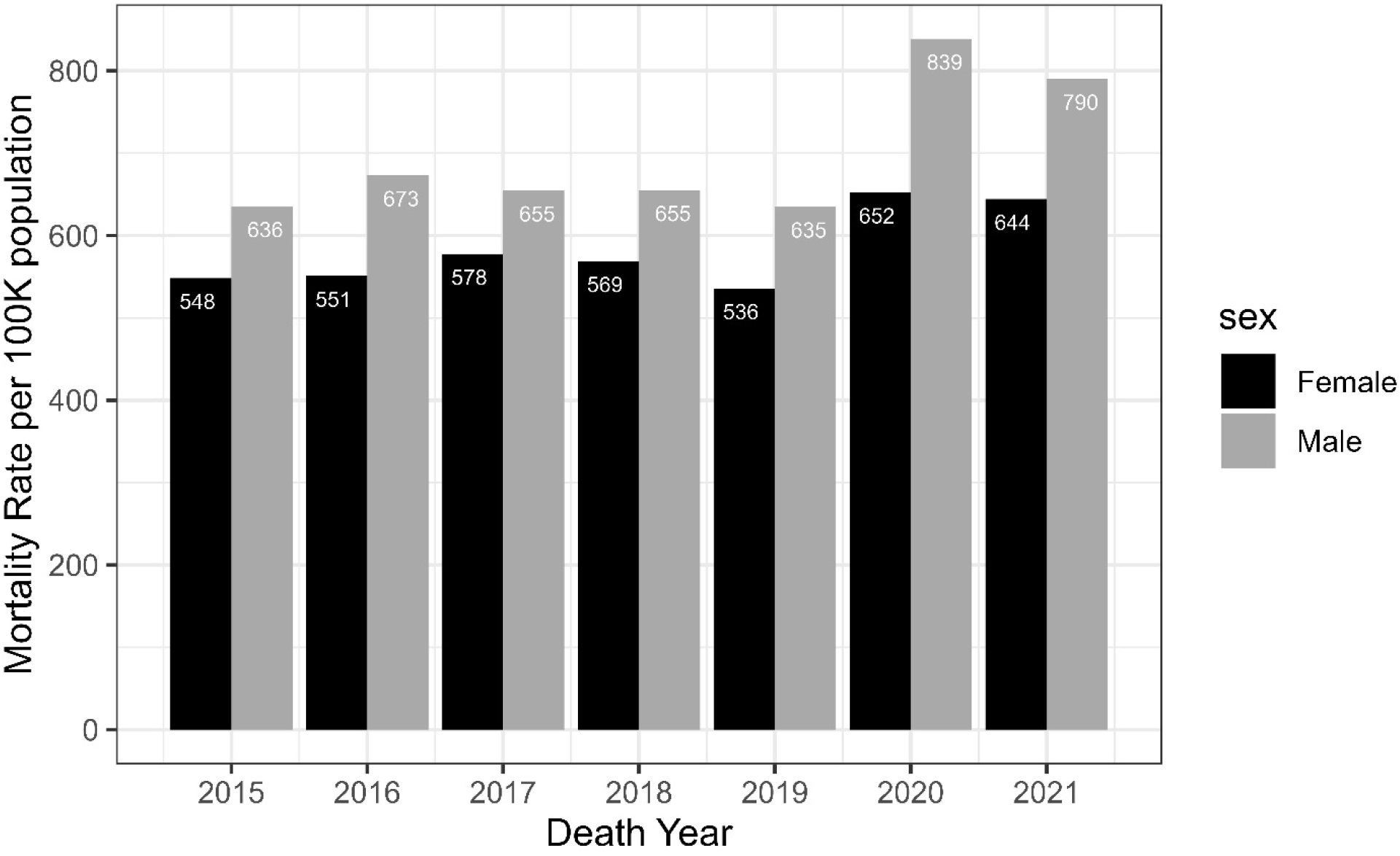
The mortality rate grew more for males than females in 2020 and 2021 in Washington, DC.

As previously argued, COVID-19 deaths can be under-reported in official statistics because people who died with other comorbidities had not been diagnosed with COVID-19. Our findings support this hypothesis: among DC residents that died in the district in 2020, there were 689 (97 per 100K) deaths where COVID-19 was listed as a cause of death, and 363 (54 per 100,000) COVID-19 deaths in 2021.

Changes in specific causes of death also reveal disparities by gender and race. **Figure 4** shows changes in the mortality rate between 2020 and the previous five years combined for some of the most common causes of death in DC by race and sex (Appendix G shows a very similar pattern for 2021, compared to the 2015-2019 period). Accidental deaths, violent deaths, suicides, as well as stroke and heart disease-related deaths grew most for Blacks (males and females), and Hispanic males. Communities disproportionately affected by poverty, systemic inequity, and structural racism incurred the heaviest loss of life due to rising gun violence. The Metropolitan Police Department reported a 34% increase in homicides in 2020 compared to the previous five-year average (29) and nationwide, and the CDC reported a 30% increase in the firearm homicide rate since the start of the pandemic (30). Blacks and Hispanics in the district are also essential and critical infrastructure workers. The U.S. Bureau of Labor Statistics reported increases in numerous construction jobs throughout 2020 (31), which might have contributed to the rise in accidents in groups with higher occupational exposure, like Black and Hispanic males. Some of the increases in mortality related to chronic conditions (e.g., heart disease, stroke COPD, and other respiratory diseases) in 2020 compared to previous years might be due to general avoidance of care linked to the perceived risk of COVID-19 exposure in healthcare facilities (7, 32) as well as the greater vulnerability of Blacks and Hispanics to chronic conditions relative to Whites.

**Figure 4.**
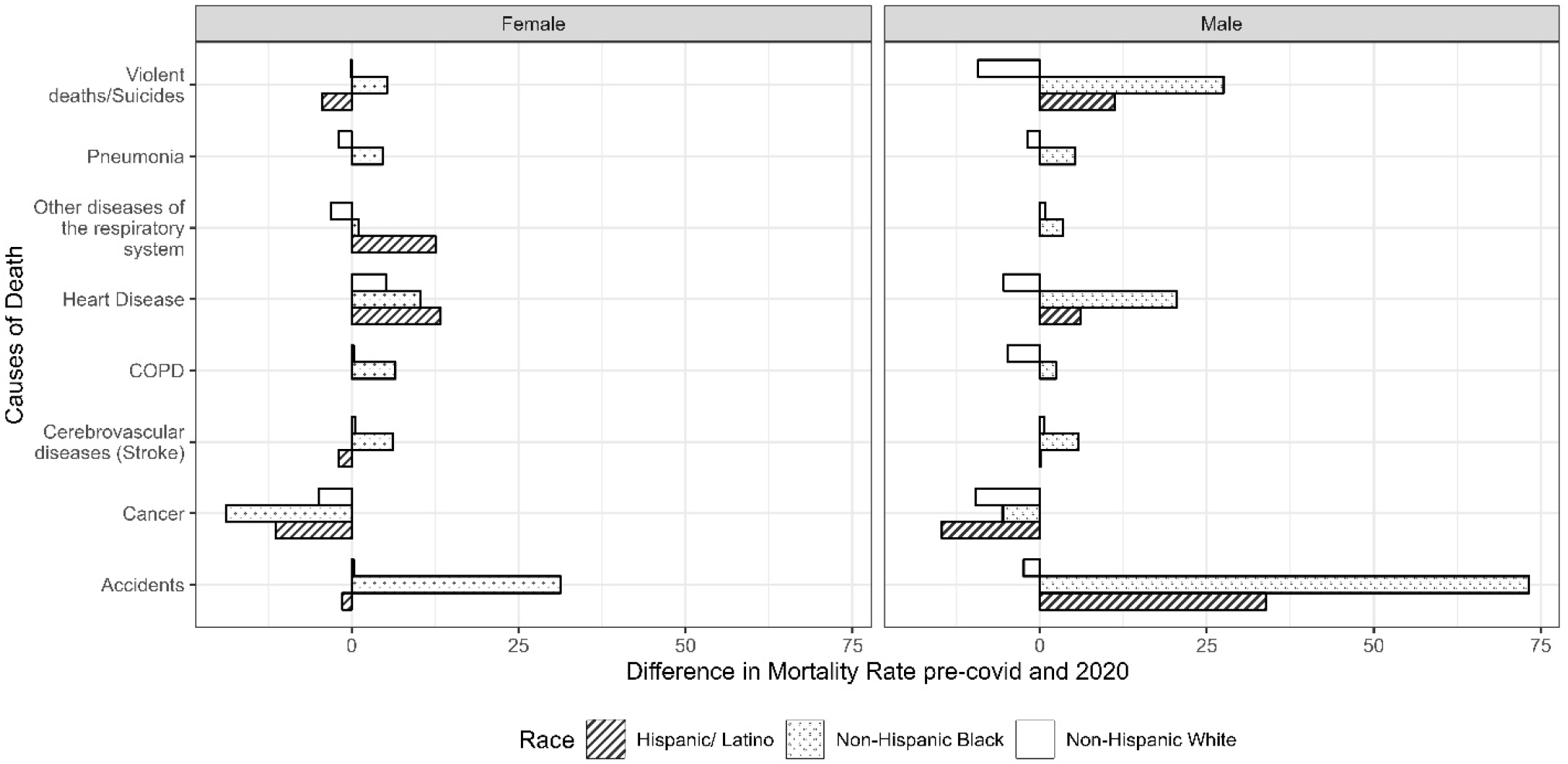
Cause-specific mortality rates grew for Blacks and Hispanics in 2020, especially for males, while rates remained flat for Whites.

The cause-specific mortality rates for Whites either decreased or stayed the same, irrespective of gender. Decreases in mortality for violent deaths and accidents among White males is likely the byproduct of lockdowns, i.e., the shift from moving out and about in the world to settling into the more protected environment of one’s home. In contrast, decreases in the mortality rate for some of the most prevalent conditions, like cancer and heart disease among the White population, might be the results of a competing risk story. That is, if White people have a relatively higher likelihood of having COVID-19 listed as the primary cause of death, reducing other cause specific death types.

### YLL

The excess deaths entailed a loss of life expectancy for the total population in 2020 and 2021, with larger decreases for Hispanics and Blacks. In 2019, the average life expectancy in Washington, D.C., was 78, 0.8 years lower than the national average (33). The national average life expectancy for Hispanic women was 84.4, for White women was 81.3 years, and for Black women was 78.1 (33). The national average life expectancy for Hispanic men was 79.1, for White men was 76.3, and for Black men was 71.3. While nationally in the first year of the pandemic we lost on average 1.5 years of life expectancy, DC lost 2.7 years, which again was unevenly distributed (Appendix H).

Using DC-specific COVID prevalence rates and relative risks of death by race and sex, we estimate that in 2020 White, Black, and Hispanic women experienced, respectively, 1.2, 2.7, and 3.1 YLL due to COVID-19. The estimated YLL for White Black, and Hispanic men in 2020 was 1.5, 3.5, and 4.5, respectively (Table1). While life expectancy improved for all in 2021—possibly due to the arrival of vaccines and the advent of less severe variants—non-Hispanic Black people continued to have higher prevalence rates than the rest of the population, and Hispanics had a much higher risk of death than any other group (Appendix C). White, Black, and Hispanic women experienced, respectively, 0.4, 1.2, and 1.3 YLL due to COVID-19 in 2021. The estimated YLL for White, Black, and Hispanic men in 2021 was 1.5, 3.5, and 4.5, respectively.

**Table 1.** Years of Life Loss during the first two years of the pandemic by race and sex in DC.

|  |  | Life Expectancy at birth (2020) |  | Years of Life Lost due to Covid-19 (2020) | Life Expectancy at birth (2021) |  | Years of Life Lost due to Covid-19 (2021) |
| --- | --- | --- | --- | --- | --- | --- | --- |
| Race | Sex | Without Covid-19 | With Covid-19 |  | Without Covid-19 | With Covid-19 |  |
| Hispanic | Female | 84.4 | 81.3 | 3.1 | 81.7 | 80.5 | 1.3 |
| Hispanic | Male | 79.1 | 74.6 | 4.5 | 75.7 | 73.9 | 1.8 |
| Non-Hispanic Black | Female | 78.1 | 75.4 | 2.7 | 76.0 | 74.7 | 1.2 |
| Non-Hispanic Black | Male | 71.3 | 67.8 | 3.5 | 68.4 | 67.1 | 1.3 |
| Non-Hispanic White | Female | 81.3 | 80.1 | 1.2 | 80.1 | 79.7 | 0.4 |
| Non-Hispanic White | Male | 76.3 | 74.8 | 1.5 | 74.9 | 74.4 | 0.5 |

## Discussion

The widening of the life expectancy gap in a high-income but already unequal city like Washington DC during COVID-19 emphasizes the importance of micro-level research for grasping the ripples of major health disruptions like the COVID-19 pandemic and supporting population health. In DC, a person’s race is highly correlated with income, and life expectancy. The YLL in the Hispanic/Latino and Black population is particularly concerning because it could have severe intergenerational consequences. The premature death of parents and caretakers has been shown, in other contexts, to contribute to social disadvantage across generations due to weathering (the chronic lack of emotional and socio-economic resources available to children following the death of a parent; (34)), and scarring (reduction in life satisfaction due to an adverse past experience like the death of a parent; (35)).

National statistics can mask marked differences at the state/city level and within states/cities across groups. In the US, we have not been consistent in reporting sex and race data on COVID-19 prevalence and deaths by location. Only 13 states report cases by sex (36) and only a handful of large metropolitan areas have reported cases by race (37–39). Such data limitations impede research, given that comparisons across geography and demographic subgroups depend on access to information on exposure, diagnosis, and population totals essential for computing rates. When compared with other states, the overall COVID health profile of DC as a metropolitan region is average (40) However, when health indicators are stratified by race and wealth (i.e., wards), demographic differences in mortality and YLL are remarkably unequal, as we show.

Documenting differences is a first step in determining the causes of gaps. The sex and race gap encountered in the first two years of the pandemic is not easily explained by any single factor. In the case of sex, biological explanations have been put forward, from the X-chromosome containing a high density of immune-related genes to men being more susceptible to infection due to higher ACE2 plasma levels than women (41). In the case of race, different health behaviors, from masking to vaccination uptake and occupational risks might also have contributed to the existing gaps (42) The burden of COVID-19 has been unevenly borne, depending on groups’ ability to self-protect by working from home, prevalence of pre-existing chronic conditions, and access to health care (43).

Our results show that the pandemic is a catalyst of inequality in that its consequences are particularly burdensome on already vulnerable groups. In DC, the burden of COVID-19 has reversed life expectancy gains of Black and Hispanic people. In the two decades prior to the pandemic, life expectancy increased by 3.9 years, 2.7, and 1.7 years for Blacks, Hispanics and Whites, respectively (44). Whites have made proportionately fewer gains in life expectancy compared to Black and Hispanics because of the increased mortality among middle-aged white Americans driven by the opioid epidemic (45). While COVID has played a role in shortening life expectancy, attributing the decline entirely to the virus would be misguided. Data from the Behavioral Risk Factor Surveillance System (BRFSS) has shown an ongoing increase in the prevalence of obesity and chronic conditions among low-income individuals in the district as well as nationally (46). By devoting resources to prevent COVID-specific and -adjacent deaths, we can reduce these dangerous societal imbalances and promote equitable population health.

Unfortunately, federal funding during the pandemic has not been equitable or commensurate with need. DC has received less funding than Delaware, Rhode Island, Vermont, and Wyoming, which have smaller populations and whose residents pay less in federal taxes (47). Hospital closures further restrict access to medical resources: Providence hospital, located in Ward 5, was closed in April 2019, and United Medical center, located in Ward 8, is set to close by the end of 2023, with many services halted since 2020, leaving no open hospitals east of the Anacostia River, which only compounds health inequities in Wards 7 and 8. Hispanic Americans remain less likely than other racial and ethnic groups – including White and Black Americans – to be insured or have a primary care provider (48). Overall, addressing the disproportionate burden of COVID-19 requires a multifaceted approach that addresses the lack of attention and resources currently being devoted to narrow the gap in life expectancy, from better representation from elected officials and community leaders who advocate for their community’s needs to better outreach for vaccinations, prevention of chronic conditions and violent deaths.

### Limitations

Our primary data source underestimates mortality rates in the city. While the denominator represents the count of residents in DC, the numerator represents only DC residents who have died in the district. Local departments of vital statistics do not have access to the death records of residents dying elsewhere. When the CDC aggregates the death counts from states and defines the total counts of deaths for each state and DC, we infer that approximately 14.4% of DC residents have died outside the state or overseas (Appendix I). Accounting for the demographic characteristics of residents that died outside DC would improve the accuracy of our estimates.

## Data Availability

All data in the present study were obtained from the Department of Vital Records in Washington, DC. These data are proprietary and will not be shared by the authors; we encourage interested parties to contact the Department of Vital Records regarding data access.

https://dchealth.dc.gov/service/request-record

## Acknowledgments

We are grateful to DC Health and the DC Vital Records Divisions for providing access to death certificates. This work was made possible by the National Institute of Allergy and Infectious Diseases of the National Institutes of Health under award number R03AI16397801.

## Author Contributions

MLA is responsible for the study concept, design, data acquisition, statistical analysis, and manuscript drafting. SSI had access to all the data in the study and is responsible for data cleaning, gathering, and statistical analysis. JH provided critical revisions to the manuscript. All authors contributed to the interpretation of data, and reviewed and approved the final manuscript for submission.

## Supplemental Materials

**Appendix A.** Total population estimates in Washington DC by year and racial breakdown

| Year | Total population<br>(% Margin of Error) | % Black<br>(% Margin of Error) | % White<br>(% Margin of Error) | % Hispanic<br>(% Margin of Error) |
| --- | --- | --- | --- | --- |
| 2015 | 672,228 (± 0.1) | 46.5% (± 0.3) | 36.0 % (± 0.1) | 10.6% (± 0.8) |
| 2016 | 681,170 (± 0.1) | 46.4% (± 0.3) | 36.3% (± 0.1) | 10.9% (± 0.7) |
| 2017 | 693,972 (± 0.1) | 45.3% (± 0.4) | 36.5% (± 0.2) | 11.0% (± 0.8) |
| 2018 | 702,455 (± 0.3) | 44.4% (± 0.4) | 36.9% (± 0.1) | 11.3% (± 0.7) |
| 2019 | 705,749 (± 0.1) | 44.1% (± 0.4) | 37.3% (± 0.1) | 11.3% (± 0.7) |
| 2020 | 712,816 (± 0.1) | 44.1% (± 0.4)* | 37.3% (± 0.1)* | 11.3% (± 0.7)* |
| 2021 | 670,050 (± 0.1) | 42.7% (±0.4) | 36.4% (±0.3) | 11.5% (±1.0) |

Source: US Census Bureau (1)
\*Note: Because the experimental data tables released by the Census for 2020(2) did not have a breakdown by race, we used the same race composition as 2020.
Margin of Error for Total population and Hispanic population estimates are calculated by aggregating the standard errors of the subgroups.
Standard Error = Margin of Error (90% confidence level)/1.645
Where Margin of Error = max (upper bound - estimate, estimate - lower bound)
Where $SE(\widehat{X}_1)$ , $SE(\widehat{X}_2)$ etc. are standard errors of sample estimates and $SE(\widehat{X}_1 + \widehat{X}_2 + \dots)$ is the approximate standard error of the sum of these sample estimates.

**Appendix B.**
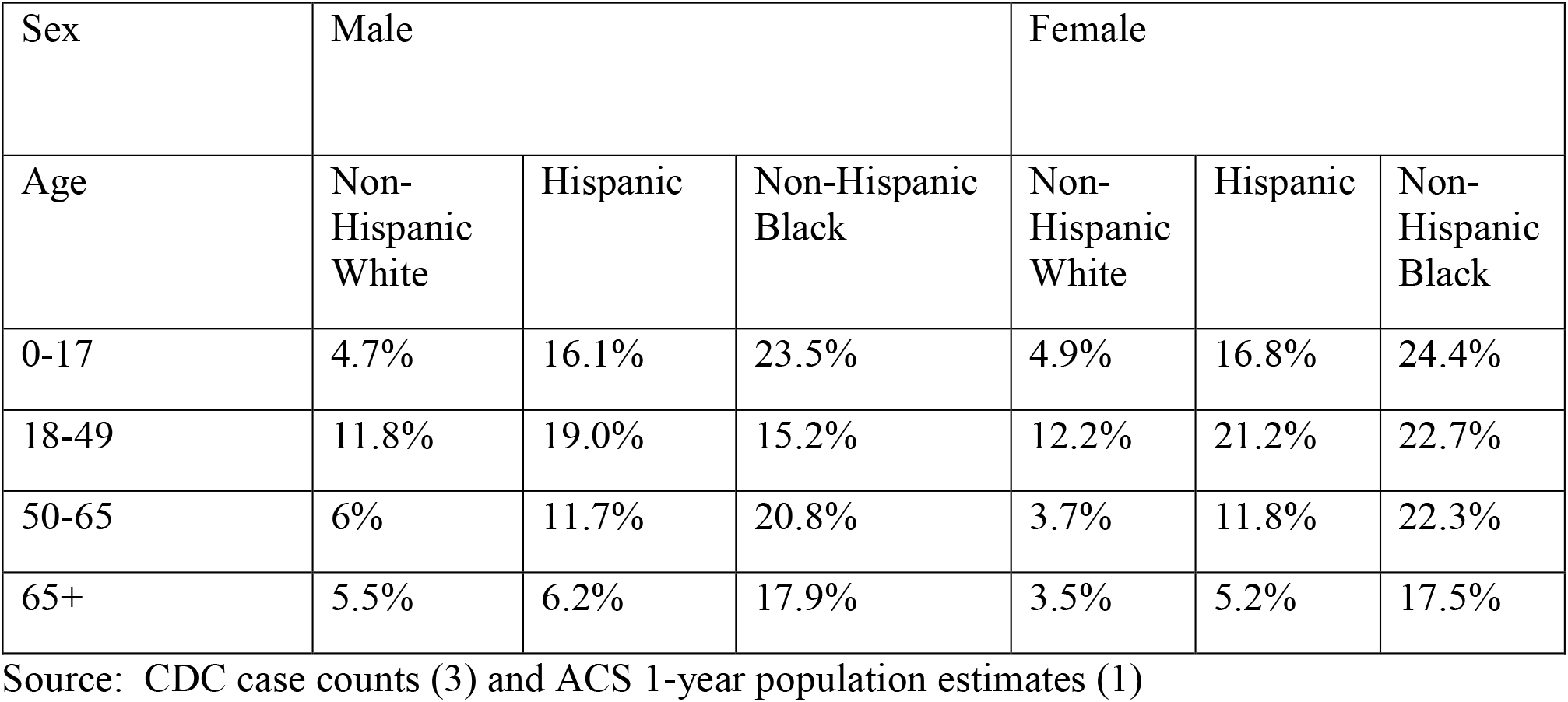
COVID-19 Prevalence Rate (%) by Age, Sex, and Race in DC, 2020-2021

**Appendix C.** Relative Risk of COVID Deaths by race in DC

| Sex | Male |  |  | Female |  |  |
| --- | --- | --- | --- | --- | --- | --- |
| Year | Non-Hispanic White | Hispanic | Non-Hispanic Black | Non-Hispanic White | Hispanic | Non-Hispanic Black |
| **2020** | 1.12 | 1.71 | 1.15 | 1.09 | 1.35 | 1.14 |
| **2021** | 1.09 | 1.16 | 1.1 | 1.04 | 1.13 | 1.1 |

Source: Author's calculations
Relative Risk calculation:
Numerator: number of people dying from COVID-19
Denominator number of people who died from causes other than COVID-19

**Appendix D.** Total population estimates by race and ward

| Ward | Population | % Black | % White | % Hispanic | Median Income |
| --- | --- | --- | --- | --- | --- |
| 1 | 85,289 | 25.4% | 46.9% | 17.6% | \$110,457 |
| 2 | 77,923 | 7.9% | 66.1% | 12.9% | \$114,649 |
| 3 | 84,651 | 8.1% | 70.4% | 9.6% | \$129,681 |
| 4 | 90,300 | 48.4% | 23.6% | 22.5% | \$96,245 |
| 5 | 92,094 | 61.3% | 22.5% | 10.4% | \$77,367 |
| 6 | 101,882 | 27.7% | 56.3% | 7.8% | \$117,752 |
| 7 | 84,271 | 90.2% | 2.7% | 4.0% | \$48,474 |
| 8 | 85,564 | 86.0% | 5.9% | 4.1% | \$37,963 |

Source: Office of Planning, Open Data DC (4)
Note: Data at the ward level is not available after 2018. Median Income is in 2020 inflation-adjusted dollars.
Note: The three racial groups account for approximately 93% of the population. The remainder represents the following groups as described by the ACS: Asians, American Indian and Alaska Native, Native Hawaiian and Other Pacific Islanders, and Some Other Race.

**Appendix E:**
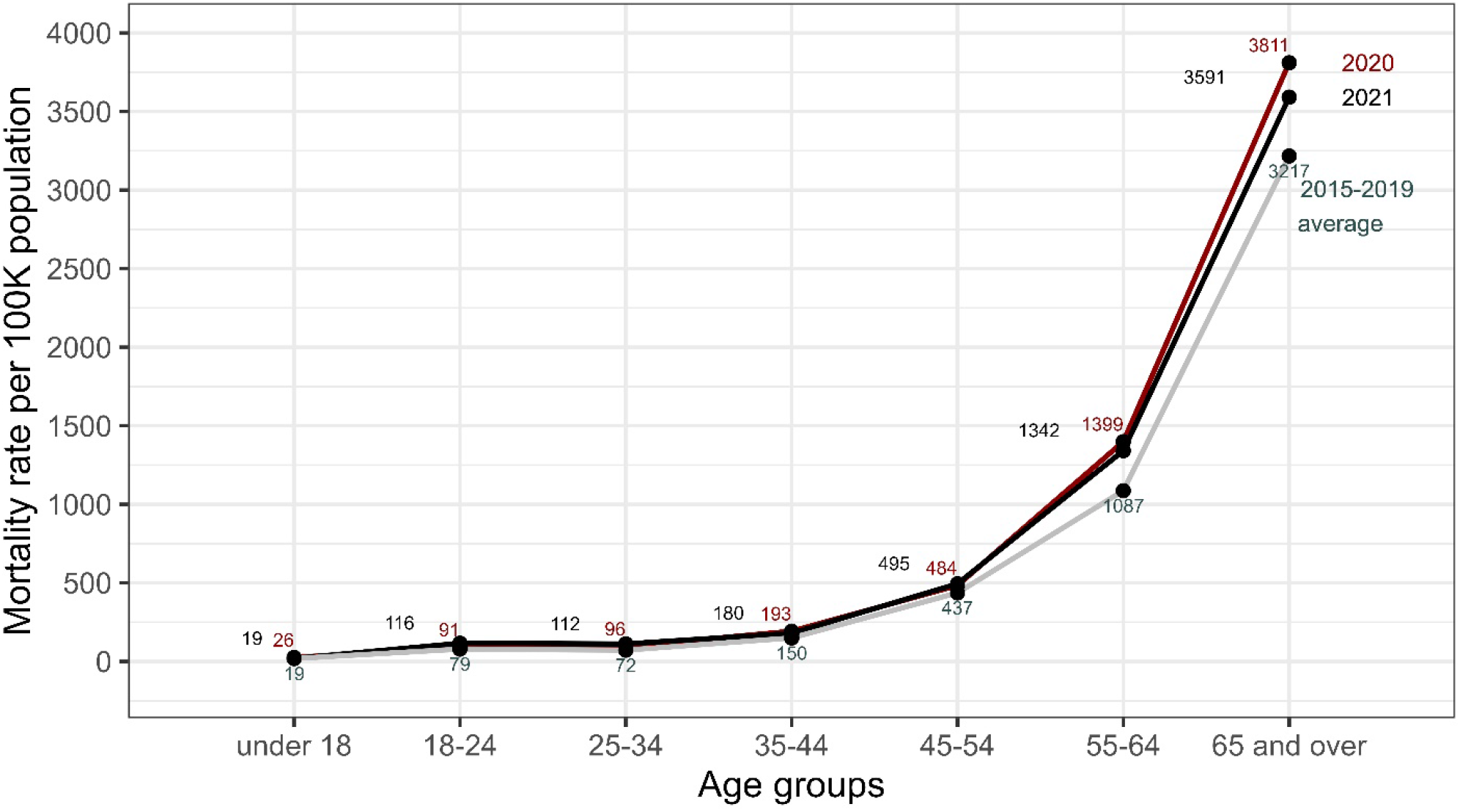
Between 2020-2021 and the previous five years (2015-2019), excess deaths show that residents 45 and older are most affected

**Appendix F:**
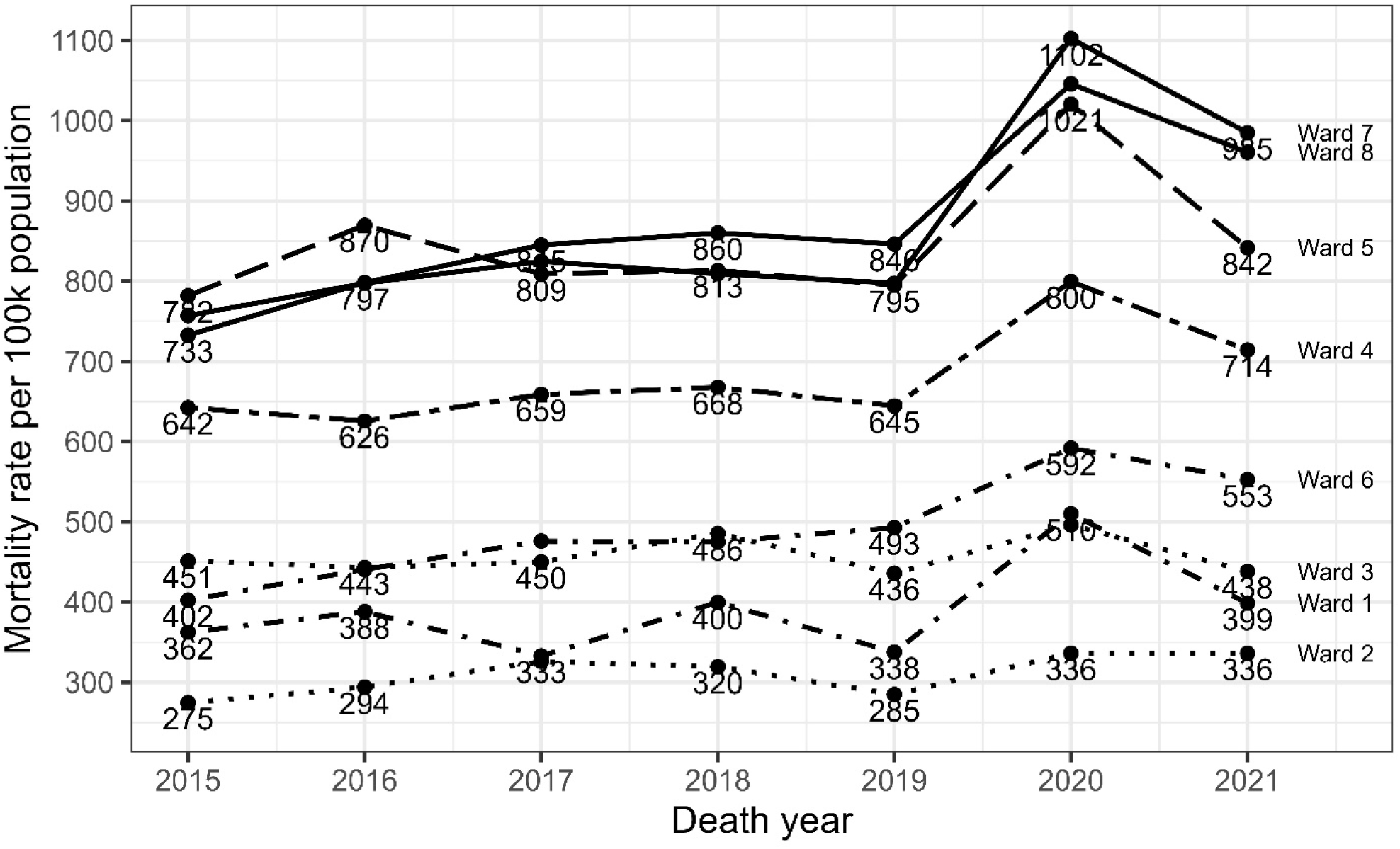
Increases in mortality rate in 2020 were greater for DC wards with more ethnic minorities and lower-income residents

**Appendix G:**
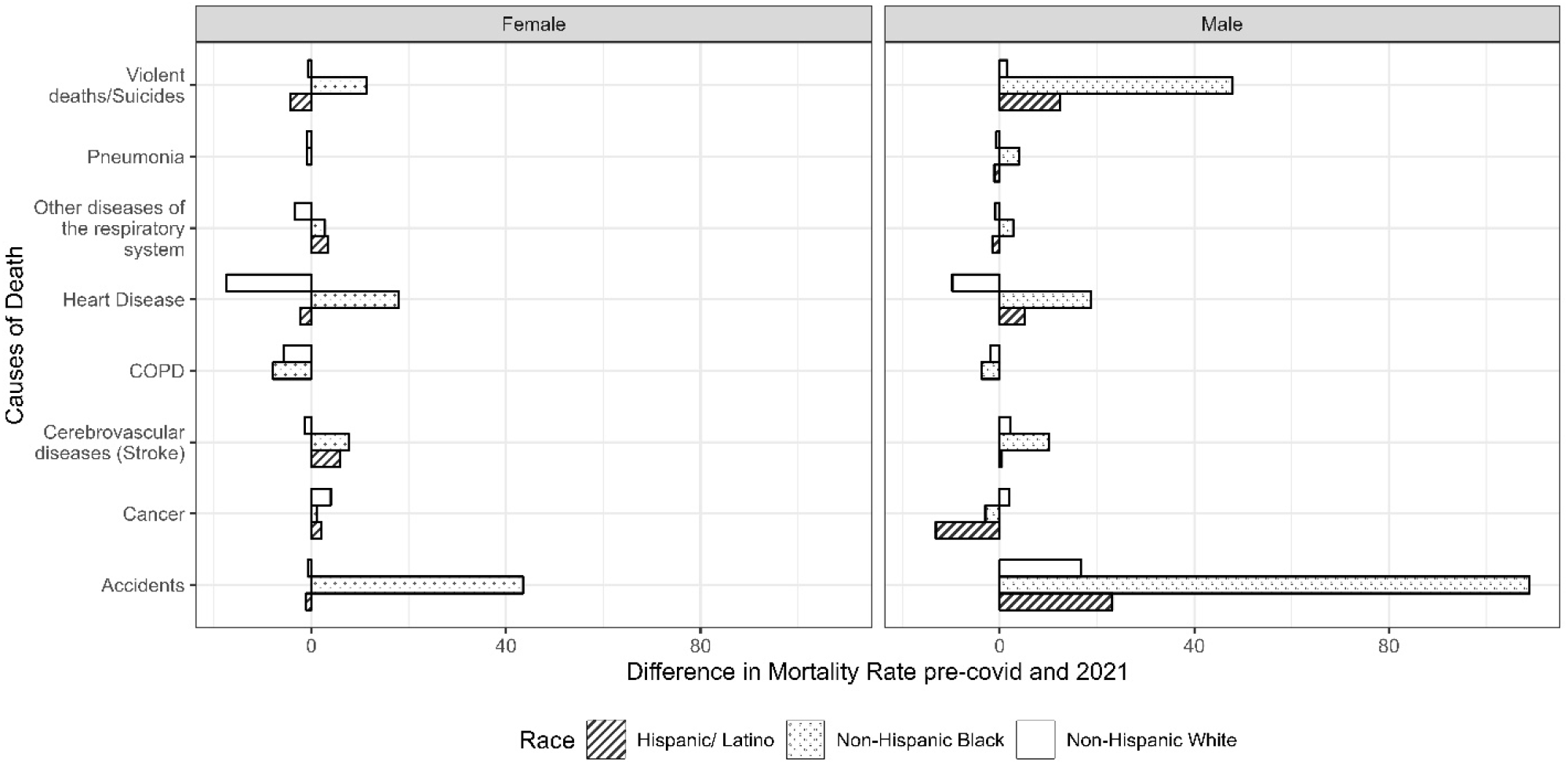
Cause-specific mortality rates by race and sex in 2021, compared to the 2015-2019 period

**Appendix H.**
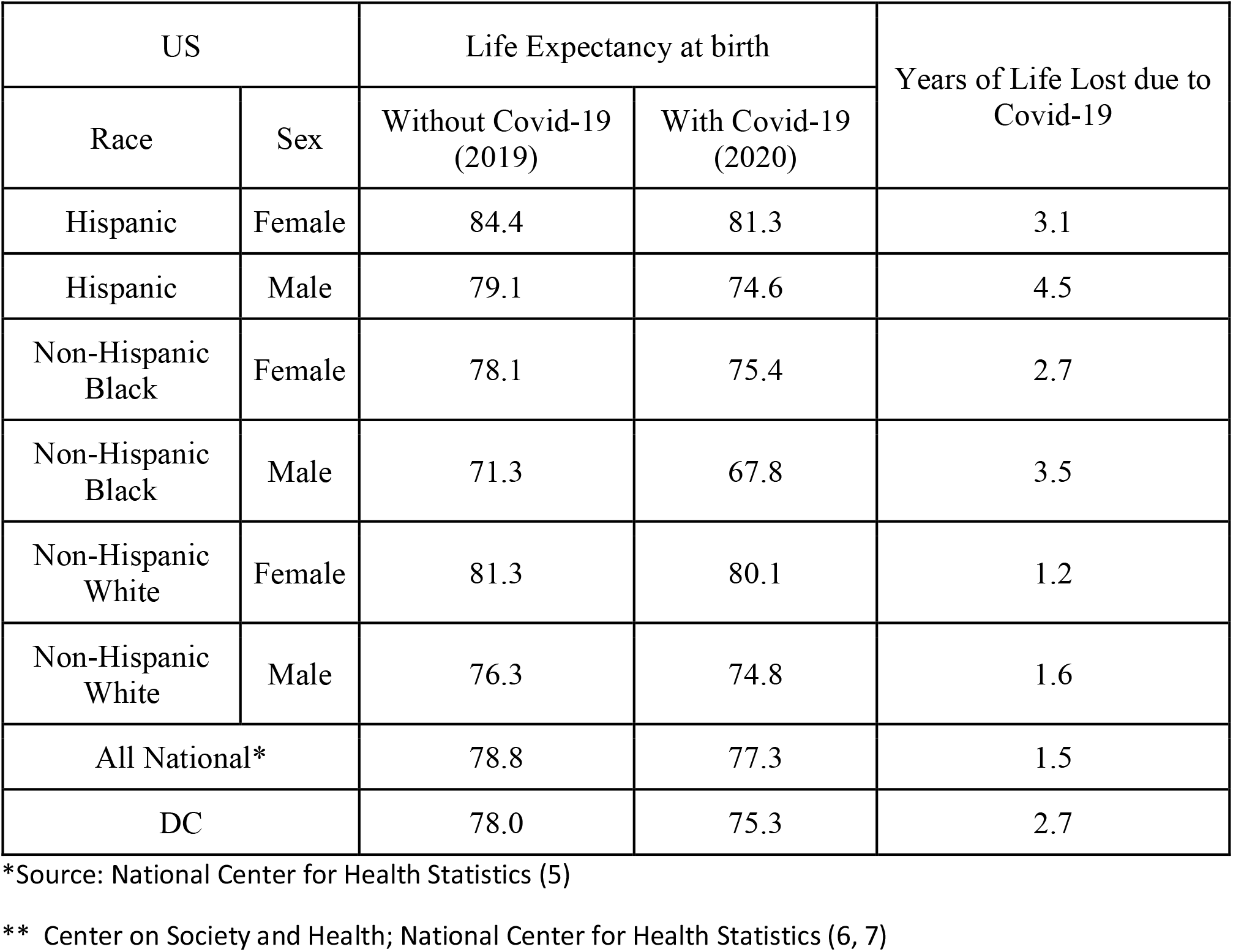
Differences in Life expectancy in the US between 2019 and 2020: DC lost nearly twice as many years of life expectancy between 2019 and 2020 compared to the US as a whole

**Appendix I.** DC Department of Vital Statistics versus CDC mortality counts

| Year | DC Department of Vital Statistics mortality | CDC Mortality (as of 05-01-2023) |
| --- | --- | --- |
| 2015 | 3965 | 4871 |
| 2016 | 4148 | 5037 |
| 2017 | 4262 | 4965 |
| 2018 | 4286 | 5008 |
| 2019 | 4115 | 4927 |
| 2020 | 5283 | 6198 |
| 2021 | 4784 | 5833 |

Sources: DC Department of Vital Statistics (8) and CDC WONDER (9)
Notes: 2020 values are still provisory for the CDC. The CDC numbers include DC residents who die in other states which translated into higher death numbers

